# Investigating the first stage of the COVID-19 pandemic in Ukraine using epidemiological and genomic data

**DOI:** 10.1101/2021.03.05.21253014

**Authors:** Yuriy Gankin, Alina Nemira, Vladimir Koniukhovskii, Gerardo Chowell, Thomas A. Weppelmann, Pavel Skums, Alexander Kirpich

## Abstract

The novel coronavirus SARS-CoV-2 was first detected in China in December 2019 and has rapidly spread around the globe. The World Health Organization declared COVID-19 a pandemic in March 2020 just three months after the introduction of the virus. Individual nations have implemented and enforced a variety of social distancing interventions to slow the virus spread, that had different degrees of success. Understanding the role of non-pharmaceutical interventions (NPIs) on COVID-19 transmission in different settings is highly important. While most such studies have focused on China, neighboring Asian counties, Western Europe, and North America, there is a scarcity of studies for Eastern Europe. The aim of this study is to contribute to filling this gap by analyzing the characteristics of the *first months of the epidemic* in Ukraine using agent-based modelling and phylodynamics. Specifically, first we studied the dynamics of COVID-19 incidence and mortality and explored the impact of epidemic NPIs. Our stochastic model suggests, that even a small delay of weeks could have increased the number of cases by up to 50%, with the potential to overwhelm hospital systems. Second, the genomic data analysis suggests that there have been multiple introductions of SARS-CoV-2 into Ukraine during the early stages of the epidemic. Our findings support the conclusion that the implemented travel restrictions may have had limited impact on the epidemic spread. Third, the basic reproduction number for the epidemic that has been estimated independently from case counts data and from genomic data suggest sustained intra-country transmissions.

## 1. Introduction

SARS-CoV-2 virus causing COVID-19 was first detected in December 2019 in the Chinese city of Wuhan [1][2][3][4][5], and has rapidly spread around the globe, prompting the World Health Organization (WHO) to declare a pandemic in March, 2020 [6], just three month after the first reported case. Despite having much lower case-fatality rate than other recent coronavirus pandemics such as the severe acute respiratory syndrome (SARS) and Middle East respiratory syndrome (MERS), the novel coronavirus has claimed more lives just within a few months of introduction than both of those epidemics combined [7]. As of June 29, 2021 there were more than 182 million infections with over 3.9 million deaths [8]. In the absence of vaccines during the early pandemic period, non-pharmaceutical interventions (specifically, non-pharmaceutical epidemic mitigation interventions) were the only tools at the disposal of public health authorities to prevent and to mitigate the virus spread [9][10][11]. The strategies implemented and enforced by governments around the world were highly variable and included frequent sanitation of public spaces, enforced social distancing, wearing of masks, closure of schools, churches, and ban of mass gatherings [12][13][14].

Even well after a year since the epidemic started, fundamental questions regarding the effects of non-pharmaceutical interventions (NPIs) [15][16] and the genomic evolution of SARS-CoV-2 [17][18][19][20] during the introductory period remain. Additionally, recent modeling efforts aimed at shedding light on those questions have mostly focused on China [1][2][3][4], the rest of Asia [21][22][23], Western and Central Europe [24][22][25], and North America [26][27][28][29][5], largely neglecting Africa, the Middle East, and Eastern Europe.

In Eastern Europe, post-socialist economics and healthcare systems are inherently different from Western Europe. The available SARS-CoV-2 transmission models for Eastern Europe are based on relatively simple *SIR* or similar compartmental models [30][31][32] where individuals are assigned to groups and all individuals within a given group are expected to have the same characteristics. To the best of our knowledge no agent-based modeling studies have been conducted for Ukraine to evaluate the impact of spatial heterogeneities in key transmission drivers such as density of infected individuals and their geographic locations. Furthermore, the number of genomic epidemiology studies on the COVID-19 pandemic in Eastern Europe has been limited. In this paper, we sought to fill the knowledge gap for the Ukrainian epidemic [33], which provides a unique setting for studying the COVID-19 spread under the ex-USSR healthcare system, and with the epidemic mitigation policies similar to the rest of Europe.

The first confirmed case in Ukraine was reported on March 3, 2020 and was an individual who has recently traveled from Italy. The first death was reported on March 13, 2020 [34][33]. The Ukrainian government started to implement quarantine measure on March 12, 2020 [33][32] while the cases continued to rise possibly because of the delayed detections of existing infections and returns of infected Ukrainians from abroad [32] (Figure 3B). As a result, more strict measures have been implemented on April 6, 2020 which included the closure of schools, universities, shopping malls, and mandatory mask regiment in public places [32]. Those measures were slightly softened on April 24, 2020 and many services resumed even though some restrictions lasted till the end of June 2020 [33]. As a summary, Ukrainian officials took the epidemic very seriously from the beginning and started to implement the mitigation efforts and corresponding regulations almost immediately after multiple cases in the country have been detected. At the same time implementation of the proposed mitigation efforts did vary from region to region, and so did the compliance with those regulations [35][36].

## 2. Methods

### 2.1. Agent-based Stochastic Model

To investigate the COVID-19 epidemic in Ukraine and to assess its dynamics under different mitigation scenarios, we utilized our general stochastic agent-based modeling framework [37]. The model was adjusted to the Ukrainian settings and fit into the observed Ukrainian data. The summary of the framework together with the adaptation details are outlined below.

In brief, the model simulates the epidemics evolving over the discrete time interval (1, …, *T*) with time points 1*≤ t ≤T* corresponding to calendar days and over the certain geographical area projected on a plain. Infected individuals are represented as agents with multiple characteristics that include geographic coordinates; age; infection time, severity and current status; disease stage; infectivity rate and infectivity radius which determines how frequently and where it produces secondary infections. The summaries of empirical reproduction numbers of individual agents which are generated by model simulations are used for the estimation of the population basic reproduction number *ℛ*_0_ [38]. The geographical part of the model includes circular local epidemic spread areas ***E*** = {*E*_1_, *E*_2_, …, *E*_*I*_ }characterized by their centers and radii. The centers of these areas represent hostspots of the infection introduction into the local population (e.g. transport hubs or administrative centers). The model incorporates NPI measures via a reduced infection transmission parameters which are effective starting from a certain calendar date customizable within the model.

In this study, we used epidemic spread areas and the corresponding incidence and mortality data reported by the National Security and Defence Council of Ukraine [39]. It includes daily reports for individual administrative regions (”oblast”) under the control of the Ukrainian government starting from March, 2020. The reported data was separated into three parts. The initially reported cases from March 12, 2020 to April 12, 2020 were retrospectively incorporated into the model as the initial conditions [37]. The reported and model-produced data from April 22, 2020 to July 12, 2020 were used for model calibration, and from July 13, 2020 to August 1, 2020 – for model validation. The data before April 22, 2020 were used solely for the initial conditions to increase the model fit robustness, since the initial number of cases was relatively small in comparison to subsequent periods. The August 1, 2020 has been selected as the end date of our simulations to agree with the dates of genomic analysis based on available analyzed SARS-CoV-2 sequences collection times [40].

Optimization of model parameters has been performed by minimizing the sum of squared differences between the model-produced outputs (across multiple runs) and the calibration data using the Nelder–Mead numerical minimization method [41]. The population basic reproduction number *ℛ*_0_ [38] has been estimated from the model-produced distribution quantiles (5%, median, 95%) of the reproduction numbers of individuals and summarized across multiple stochastic runs [37]. The estimates for *ℛ*_0_ were produced from the model fit to real data with the assumption that interventions have started almost immediately after the virus introduction.

In addition to the simulations based on the model fit to the actual case count, mortality and NPI data, two alternative simulation scenarios were considered under the hypothetical assumptions that NPIs that caused reduced transmissibility were implemented one (on April 19, 2020) and two (on April 26, 2020) weeks after the simulation start time. The results of simulations under these three scenarios were compared to assess the effect of timely NPI implementations.

The additional details about the model can be found in our earlier study [37], and the model implementation tailored to Ukrainian data is available at https://github.com/quantori/COVID19-Ukraine-Transmission.

### 2.2. Genomic Epidemiology Analysis

Sixty high-quality SARS-CoV-2 genomes from Ukraine sampled between April 24, 2020 and August 7, 2020 were extracted from GISAID [40]. These genomes were utilized to construct a maximum likelihood phylogeny using Nextstrain build for SARS-CoV-2 with the default country-specific subsampling settings [42]. The obtained timed phylogeny contained Ukrainian sequences together with a representative subsample of 6479 sequences from other geographic regions, and included inferred ancestral geographic traits of internal nodes. Using these traits, intra-country transmission clusters were identified as clades with the most recent common ancestors (MRCA) estimated as originating from Ukraine. For each cluster, confidence intervals for emergence times for MRCA and its parent were also obtained.

Next, a phylodynamic analysis of the three largest clusters and the entire Ukrainian SARS-CoV-2 population was performed using BEAST v1.10.4 [43]. We used a strict molecular clock, HKY+Γ nucleotide substitution model, a tree prior with exponential growth coalescent. Priors for the parameters were defined in BEAUti v 1.10.4 and were the following: a) normal *𝒩* (*mean* = 8.0e-4, *st*.*dev* = 2.0e-5) for the clock rate, b) log-normal *ℒ𝒩* (*mean* = 1.0, *st*.*dev* = 1.25) for the population size, c) double exponential (Laplace) distribution *𝒟ℰ𝒳𝒫*(*µ* = 0, *b* = 100) for the growth rate, d) normal *𝒩* (*mean* = 0, *st*.*dev* = 1) for the freqParameter, e) exponential *ℰ𝒳𝒫*(*mean* = 0.5, *offset* = 0) for the gammaShape parameter, and f) log-normal *ℒ𝒩*(*mean* = 1.0, *st*.*dev* = 1.25) for the kappa parameter. The detailed parameters file is available in XML format at https://github.com/alanira/COVID19-Ukraine-phylodynamics. The parameters were estimated after 30,000,000 iterations of Markov Chain Monte Carlo (MCMC) sampling, with the initial 10% values discarded as burn-in. The results were accepted if the effective sample sizes were above 200 for all parameters. The estimated exponential growth rates were used to calculate the basic reproduction numbers *ℛ*_0_ under the assumption that SARS-CoV-2 generation intervals (i.e. times between infection onset and onward transmission) were gamma-distributed [44]. We used the formula

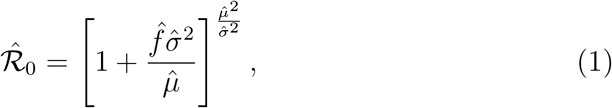

where 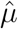 and 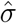 are the mean and standard deviation of the aforementioned gamma distribution [45][46][47][48]. For these values, we used the estimates 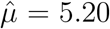 and 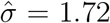 from [49] and 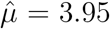and 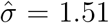 from [49]. The formula (1) defines a strictly monotone transformation of 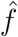, and, therefore, it also straightforwardly transforms the 95% highest posterior density (HPD) intervals for *f* into those for *ℛ*_0_.

## 3. Results

### 3.1. Agent-based Stochastic Model

The visual results of the first scenario (model fit) and the corresponding outputs are summarized in Figure 1. Blue curves in Figure 1 correspond to the reported data. They are captured by the model fits which is also indicated by the corresponding median and 90% pointwise model prediction bands across five hundred runs. The calibration interval is highlighted by cyan background.

**Figure 1:**
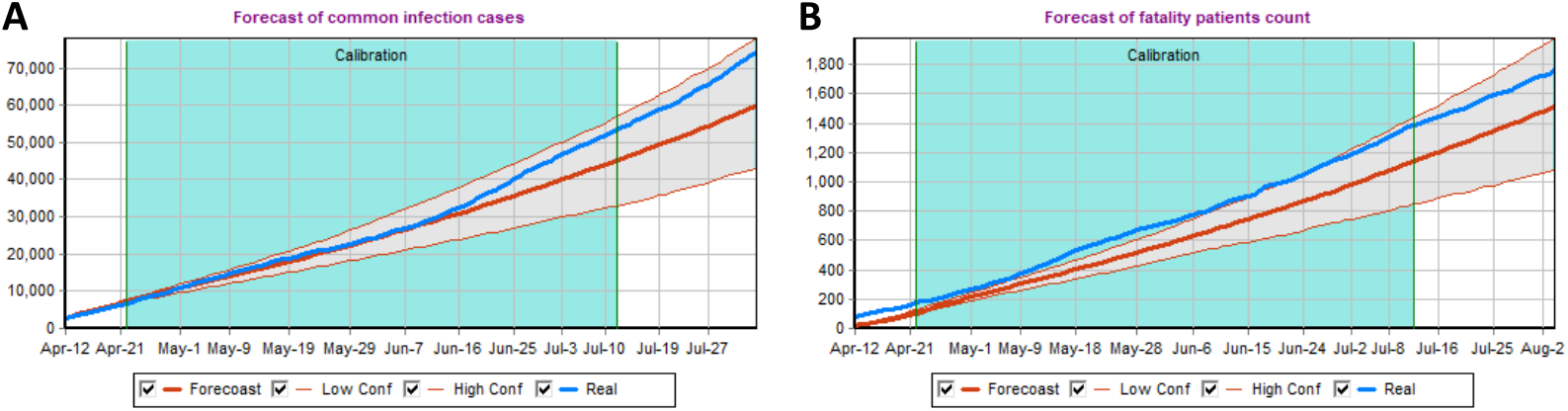
The model predictions together with the reported cases (Panel A) and reported death (Panel B) are presented. The model calibration time interval is highlighted in green. Red lines correspond to the median of the five hundred model-produced runs together with the corresponding 90% prediction bands to quantify the uncertainty. The actual observed case and death counts are displayed in blue for visual comparison.

For each of the three considered scenarios the median value across five hundred simulations were computed at each time point and presented together with the corresponding 5-th and 95-th percentiles across five hundred stochastic realizations to form the 90% prediction intervals (PI-s). The corresponding results are summarized in Tables 1 and 2 for the model-predicted cases and deaths, respectively. The three scenario summaries from Table 1 can be directly compared. For comparison the actual number of reported cases by August 1, 2020 was 71, 056 [39] which validates the model fit since August 1, 2020 was outside of the calibration interval. The hypothetical April 19th and April 26th intervention start dates produce larger number of cases in comparison to the original fitted scenario. The median estimates can be compared directly. The hypothetical April 19th scenario results in 16% predicted increase in cumulative number of cases on June 1, 2020 and in 20% predicted increase in cumulative number of cases on August 1, 2020 in comparison to the fitted scenario. The hypothetical April 26th scenario results in 36% predicted increase in cumulative number of cases on June 1, 2020 and in 46% predicted increase in cumulative number of cases on August 1, 2020 when compared to the fitted scenario.

**Table 1:** The model outputs are presented together with the reported data. The predicted number of cumulative cases produced by the model over time for three different epidemic mitigation scenarios for three initiation dates together with the corresponding 90% prediction intervals.

| Scenario | June 1, 2020 | July 1, 2020 | August 1, 2020 |
| --- | --- | --- | --- |
| Real Data | 24,012 | 44,998 | 71,056 |
| April 12, 2020 | 23,724 (19,093; 28,250) | 38,932 (29,181; 48,782) | 57,810 (41,864; 75,029) |
| April 19, 2020 | 27,511 (22,681; 32,872) | 46,193 (36,238; 57,100) | 69,358 (50,642; 88,407) |
| April 26, 2020 | 32,220 (26,016; 38,645) | 55,141 (41,974; 69,992) | 84,227 (63,992; 109,069) |

**Table 2:** The model outputs are presented together with the reported data. The predicted number of cumulative death produced by the model over time for three different epidemic mitigation scenarios for three initiation dates together with the corresponding 90% prediction intervals.

| Scenario | June 1, 2020 | July 1, 2020 | August 1, 2020 |
| --- | --- | --- | --- |
| Real Data | 718 | 1,173 | 1,709 |
| April 12, 2020 | 568 (464; 667) | 967 (738; 1,203) | 1,461 (1,050; 1,904) |
| April 19, 2020 | 653 (541; 783) | 1,143 (889; 1,412) | 1,762 (1,308; 2,241) |
| April 26, 2020 | 752 (612; 906) | 1,364 (1,049; 1,705) | 2,141 (1,604; 2,719) |

The hypothetical April 19th scenario results in 14% increase in cumulative number of deaths predicted on June 1, 2020 and in 20% increase in cumulative number of deaths predicted on August 1, 2020. The hypothetical April 26th scenario results in 32% increase in cumulative number of deaths predicted on June 1, 2020 and in 46% increase in cumulative number of deaths predicted on August 1, 2020.

Interestingly, the median results for the hypothetical April 19th scenario displayed better alignment with the actual data. This suggests the delayed impact of NPIs in transmission mitigation caused by the time needed to put the prescribed measures into effect. Furthermore, the obtained results demonstrate the importance of the early epidemic mitigation measures which cause the reduction in transmission probability parameters and, therefore, a reduction in the number of cases, and (more importantly) deaths. At the same time the results for later mitigation efforts implementation dates should only be interpreted as sensitivity analysis, since the Ukrainian government has implemented quarantine measures from the beginning of the epidemic and there were no data to properly estimate the corresponding non-intervention transmission probability parameters [50]. Therefore, the corresponding non-quarantine probability parameters have been adopted from the previous analysis [37].

The population basic reproduction number *ℛ*_0_. estimate during the intervention was estimated to be 1.10 (median) with the corresponding 90% confidence interval from quantiles equal to (0.24; 1.88).

### 3.2. Genomic epidemiology of SARS-CoV-2

Despite a sparse sampling, the observed genomic diversity of SARS-CoV-2 in Ukraine is substantial, indicating both multiple introductions of the virus and sustained intra-country evolution (Figure 2). This agrees well with the patterns observed in other countries [51], and emphasizes the contribution of global movement of people to the rapid spread of SARS-CoV-2. Specifically, Ukrainian sequences are distributed among eight lineages by the classification of [52] as follows: *B*.1 - 50.0 % of genomes, *B*.1.1 - 28.3 %, *B*1.1.243 - 8.3 %, *B*.1.527 - 5.0 %, *B*.1.1.325 - 3.3 %, and 1.7 % for each *B*.1.131, *B*.1.1.194, *B*. Similarly, by Nextstrain classification the distribution of lineages is: 19A - 1.7 %, 20A - 51.7 % and 20B - 46.7 % (Figure 2).

**Figure 2:**
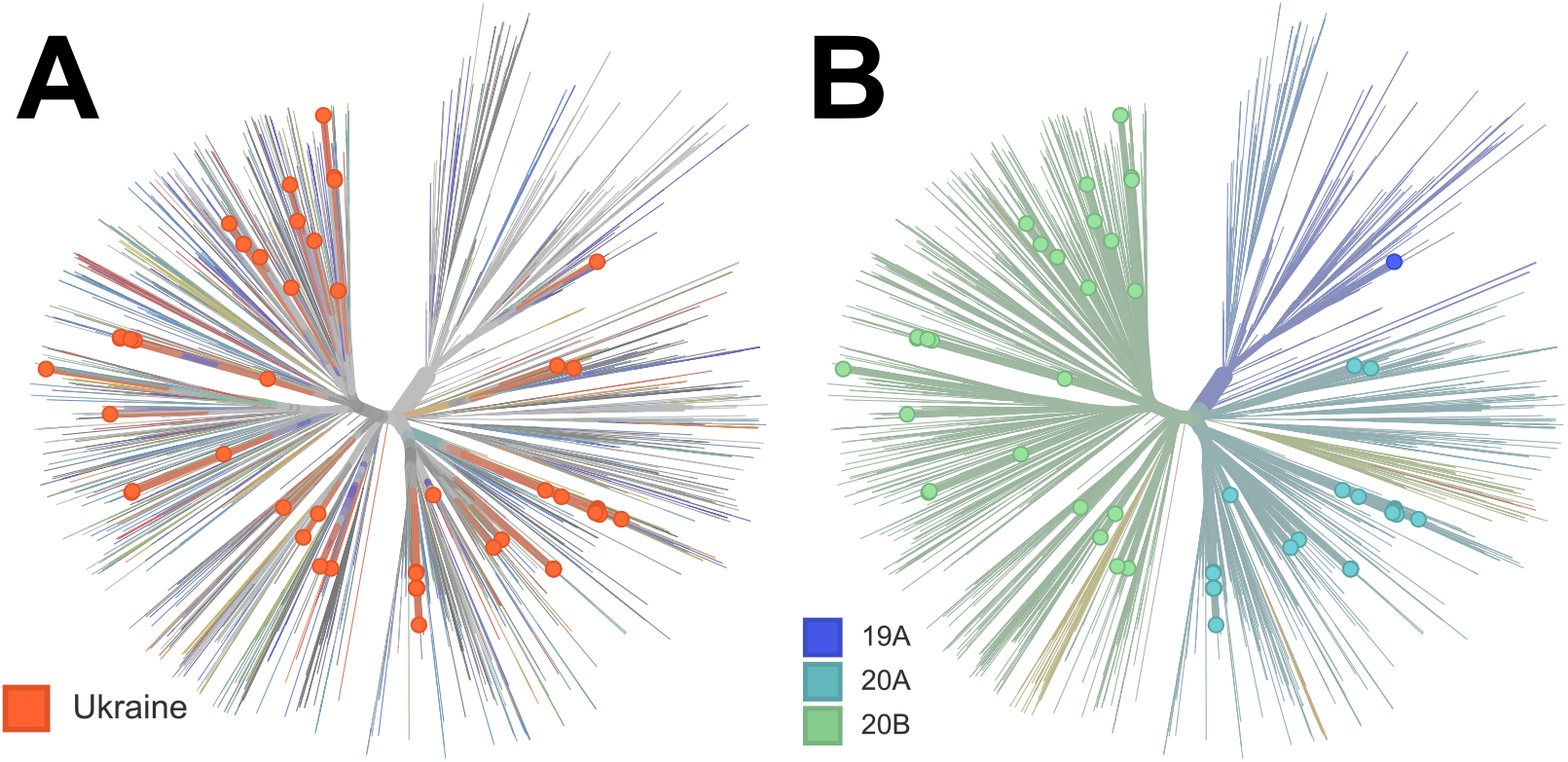
The global phylogenetic tree of SARS-CoV-2 genomes: A) distribution of Ukrainian SARS-CoV-2 genomes inside global SARS-CoV-2 population, B) the same tree with global SARS-CoV-2 lineages highlighted.

Seven Ukrainian clusters contain multiple sequences and jointly constitute 73.3% of all sampled genomes. Presence of these clusters and the corresponding intra-country lineages indicate sustained internal transmissions (Figure 3A and Figure A1-A7 in Appendix). For each such lineage, a confidence interval of its introduction time can be assessed by the union of the confidence intervals for inferred dates of its Ukrainian MRCA *v* and the non-Ukrainian parent of *v* (Table A2 in Appendix).

**Figure 3:**
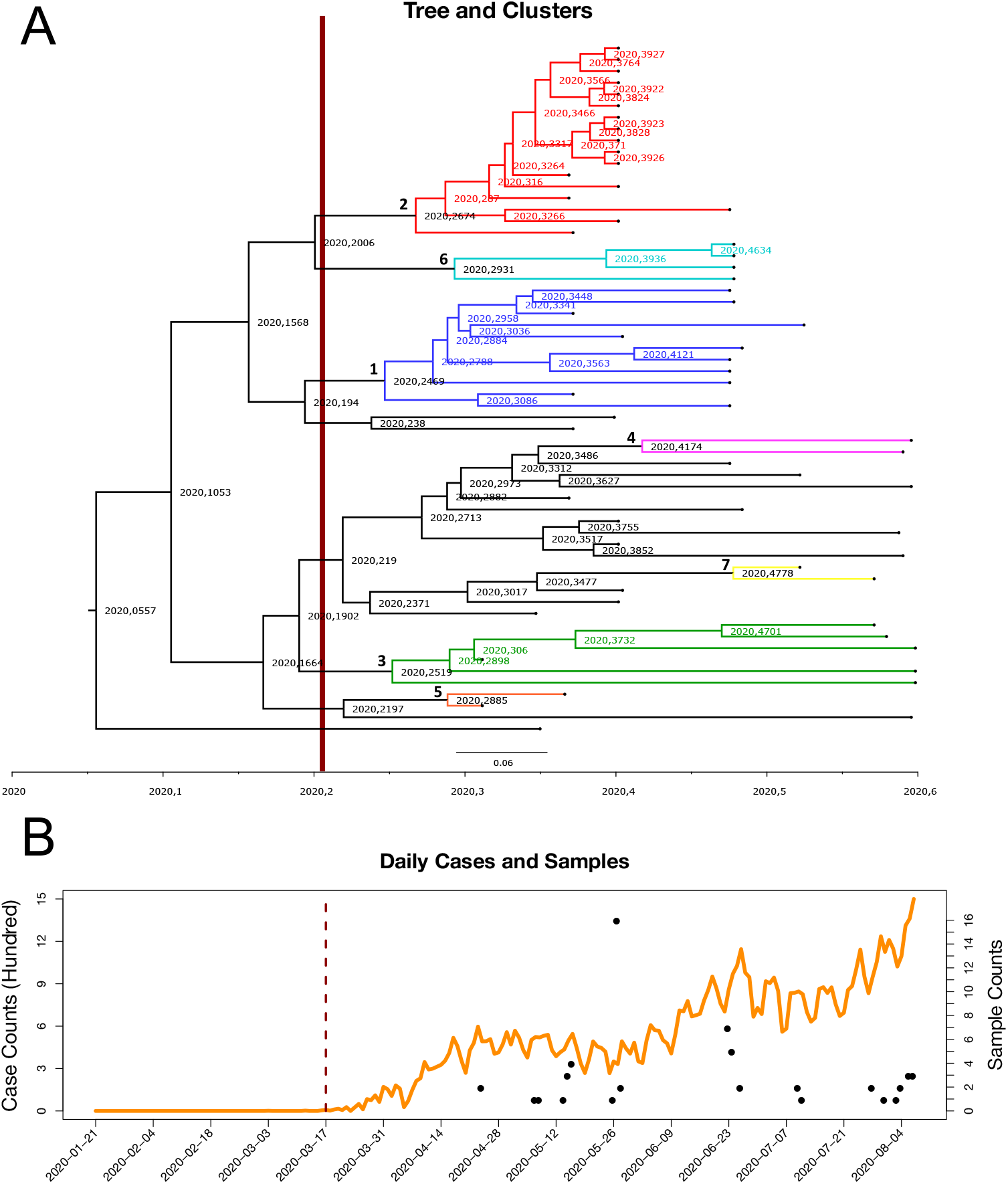
Panel A: The SARS-CoV-2 clusters are presented in the Ukrainian phylogenetic tree. Clusters colored by blue, red, green, pink, orange, azure, yellow, and numbered from one to seven, respectively. Panel B: Daily incidence of reported cases for Ukraine (orange) together with the sample counts and collection dates for sequenced samples (black). The travel restriction has happened on March 17, 2020, which is indicated by a vertical dark red bar time separator in both panels.

We analyzed these introduction times relatively to the implementation time of the travel ban, that was established on March 16, 2020 [53] for foreign citizens and on March 17, 2020 [54] for all travelers with the exception of Ukrainian citizens returning from abroad. It turned out that three out of seven lineages were most likely introduced into the country after the travel ban date, as indicated by their introduction confidence intervals (Figure 4). Similarly, a single lineage was likely imported before that date; for three remaining lineages the travel ban date falls into their confidence intervals, preventing us from the decisive conclusion, even though the date lies closer to the left ends of all intervals. Thus, the analysis support the hypothesis that the travel restrictions had limited effect on the virus importation control.

**Figure 4:**
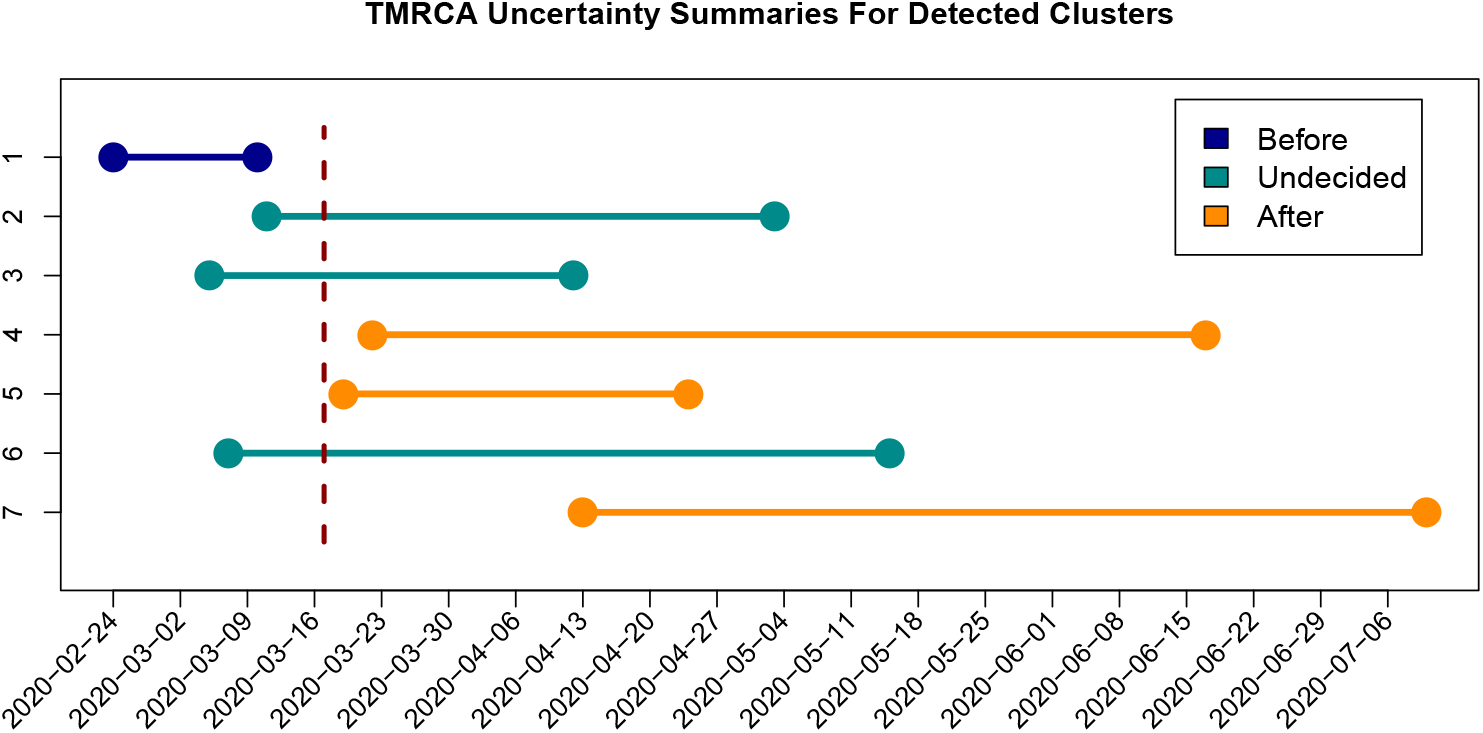
Introduction times for seven largest Ukrainian transmission lineages visualized from Table A2 from the supplement. The estimated intervals for introduction times are depicted as horizontal lines, the border closure date (March 17, 2020) is indicated by a vertical line.

The estimates of the basic reproduction number *R*_0_ for three largest lineages are summarized in Table 3. All estimates are significantly above one, indicating sustained local transmission of SARS-CoV-2 during the first months of the epidemic in Ukraine.

**Table 3:** The estimates of the basic reproduction number*ℛ* _0_ for three largest clusters together with the corresponding 95% confidence intervals (CI-s). The results are reported for two pairs of generation interval distribution parameters 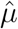 and 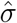 reported by two studies

| Cluster | $\hat{\mu}$ | $\hat{\sigma}$ | $\hat{\mathcal{R}}_0$ & 95% CI | $\hat{\mu}$ | $\hat{\sigma}$ | $\hat{\mathcal{R}}_0$ & 95% CI |
| --- | --- | --- | --- | --- | --- | --- |
| First | 5.20 | 1.72 | 1.31 (1.12; 1.52) | 3.95 | 1.51 | 1.23 (1.09; 1.37) |
| Second | 5.20 | 1.72 | 1.47 (1.1; 1.98) | 3.95 | 1.51 | 1.34 (1.07; 1.68) |
| Third | 5.20 | 1.72 | 1.48 (1.16; 1.94) | 3.95 | 1.51 | 1.35 (1.12; 1.65) |

## 4. Discussion

In this study, we have detailed the epidemic characteristics of the first months of the COVID-19 pandemic in Ukraine and studied the effects of NPIs. We considered two complementary approaches based on the stochastic modeling applied to incidence data and genomic epidemiology methods applied to sequencing data. Different types of data reflect various aspects of the epidemics, and are prone to different biases. Therefore, such synthetic approach facilitates ubiquitous understanding of the early stages of the epidemic in Ukraine.

COVID-19 pandemic is characterized by a richness of available data, that allow to utilize agent-based modelling and genomic analysis at the finest possible resolution. In Ukraine, we have an access to public health data on the level of individual regions, which makes agent-based model predictions more comprehensive. Similarly, the advances and cost reduction of next-generation sequencing (NGS) methods allowed rapid genomic data acquisition at early stages of the epidemic [40]. These data processed by advanced phylogenetic and phylodynamic models allow to assess the virus importation and intra-country transmission dynamics from a “different angle” [55]. Furthermore, in cases when two methods produced independent estimations of the basic reproduction number *ℛ*_0_, the obtained results are comparable, thus highlighting their consistency. The uncertainty estimates for the stochastic estimates are wider, which may be due to the fact that stochastic model has more parameters and higher variability in the outputs while phylodynamic models has pretty strong priors.

The study has limitations since the available surveillance incidence and genomic molecular data are limited. Ukraine is one of the poorest countries in Europe (based on GDP per capita), and, therefore, the health care infrastructure in Ukraine lacks in some parts the resources of its close and distant European neighbors [56]. As a result, both availability of screening tests and the reporting of incidence data during the initial epidemic likely (substantially) underestimated the burden of disease in terms of incidence counts. Likewise, as the pandemic evolved, the scarcity of genetic sequencing limited the number of sequence comparison in the phylogenetic analysis. As such, the actual number of viral clusters of local transmission remains unknown and should be interpreted as *at least seven* clusters which only form the “tip of the iceberg” of all transmission clusters. Moreover, the local population compliance with the NPI regulations implemented by officials is always a question, which might have reduced the effectiveness of such measures [35][36].

In summary, this study was among the first to explore the characteristics of the initial pandemic as it spread to Ukraine *and* provided additional *genomic analysis* not previously published.

## Data Availability

The data sources are public and are cited in the manuscript.

## 5. Acknowledgements

The authors would like to acknowledge the authors, originating and submitting laboratories of the sequences from GISAID. The list of laboratories is provided in Appendix. The authors would also like to acknowledge Sergey Brutsky for his help with Ukraine incidence data collection and acquisition.

