## Supplement for "Investigating the first stage of the COVID-19 pandemic in Ukraine using epidemiological and genomic data"

---

**Abstract**

This is the supplementary information that was not included in the main manuscript body.

---

*Equal Contribution
**Equal Contribution
Corresponding Author: Alexander Kirpich

*Preprint submitted to a research journal* *August 5, 2021*

### Appendix

*The Ukrainian Transmission Clusters*

The visualizations of seven largest intra-Ukrainian transmission clusters are presented in Figures A1-A7. The numbering of clusters corresponds to the one presented in Figures 3 and 4.

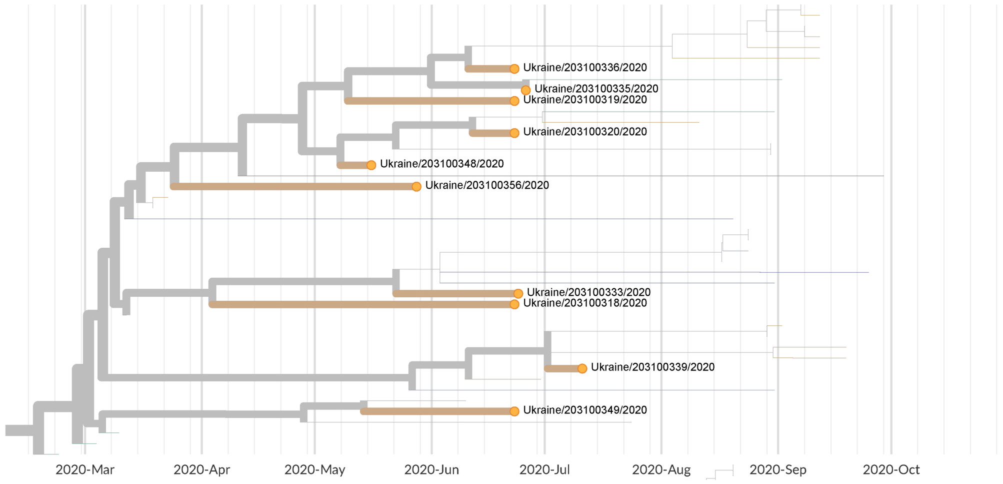


Figure A1: Cluster 1.

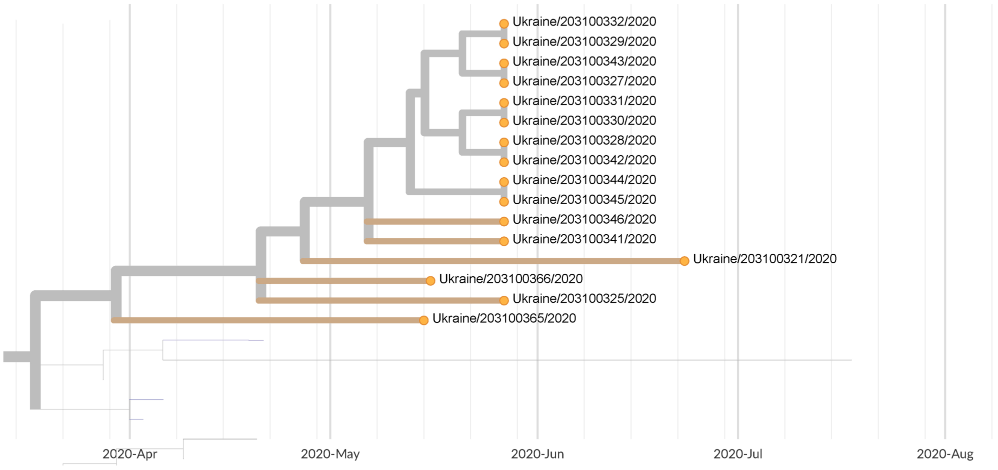


Figure A2: Cluster 2.

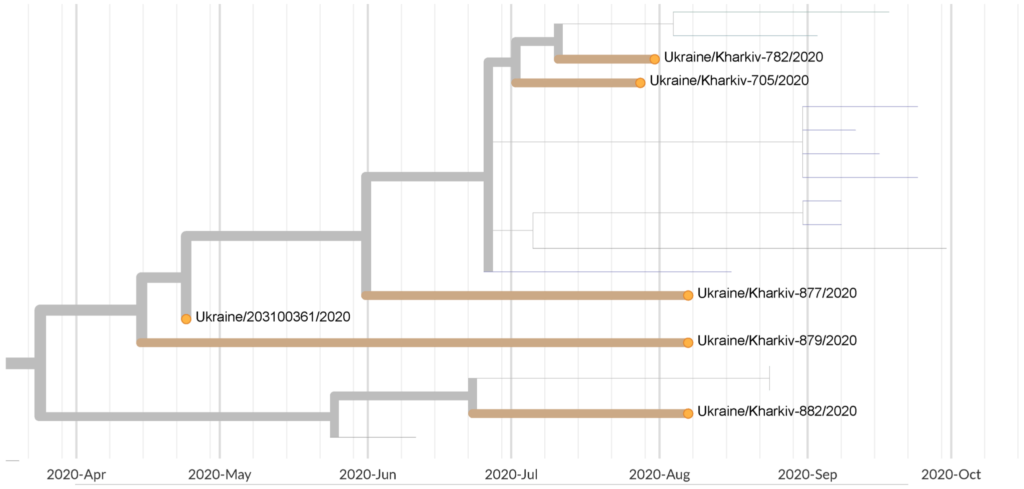


Figure A3: Cluster 3.

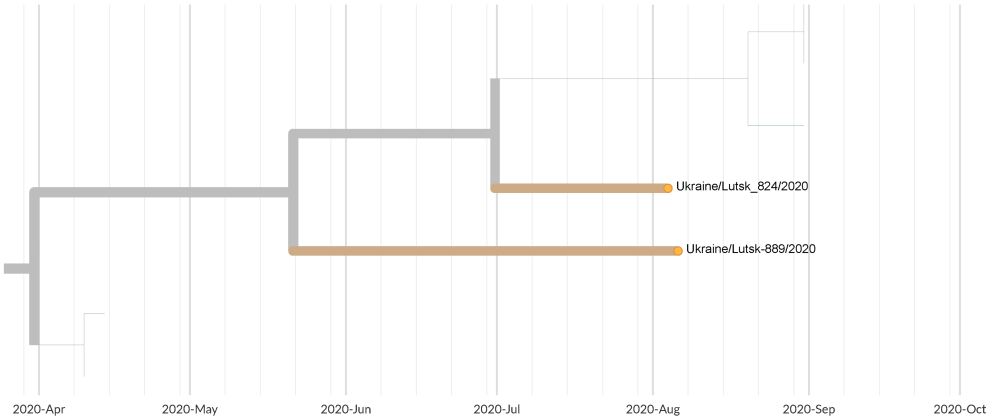


Figure A4: Cluster 4.

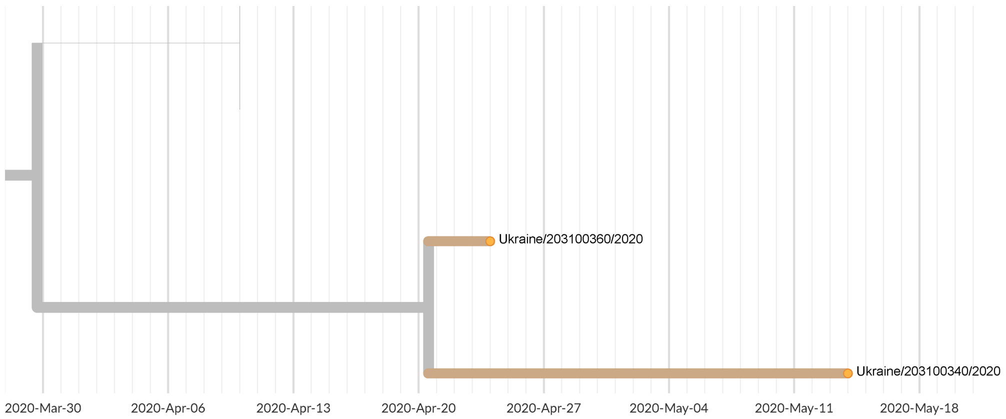


Figure A5: Cluster 5.

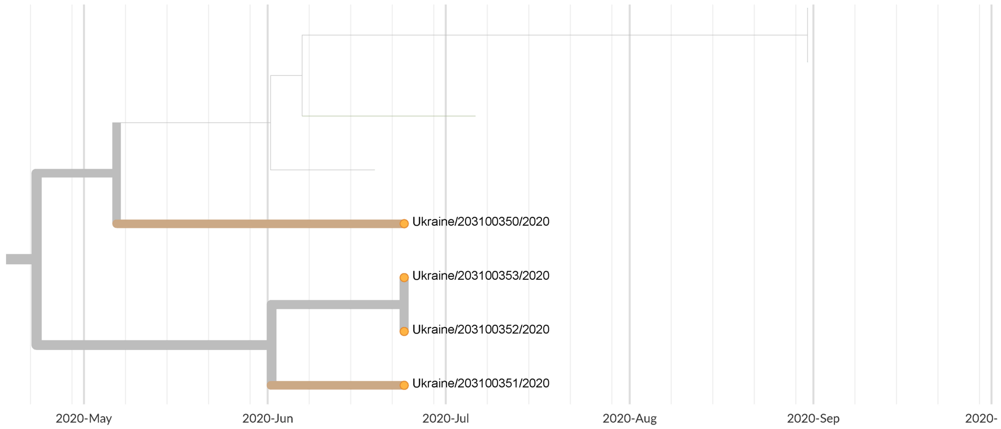


Figure A6: Cluster 6.

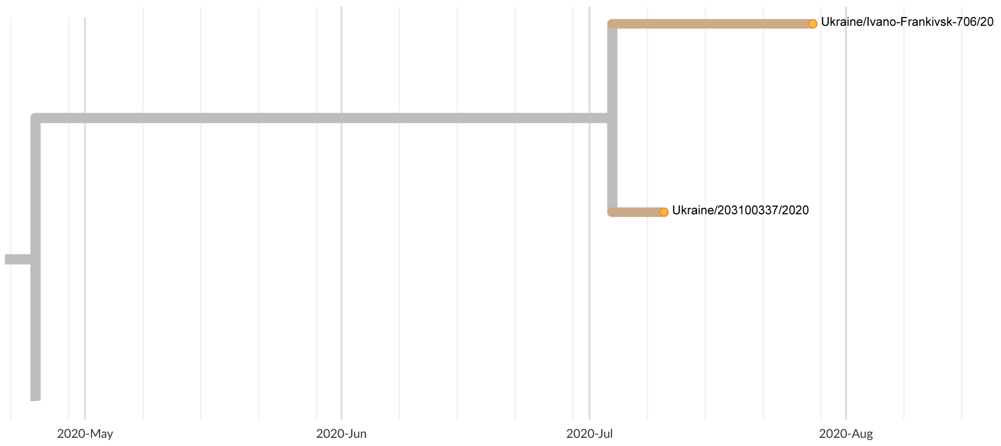


Figure A7: Cluster 7.

*TMRCAs*

The TMRCAs have been produced using Nextstrain together with the corresponding 95% confidence intervals. The results are presented in Table A2.

| **Name** | **TMRCA & CI** | **Parent TMRCA & CI** | **March 17** |
|---|---|---|---|
| Cluster 1 | 2020-03-06 (2020-02-29; 2020-03-10) | 2020-03-02 (2020-02-24; 2020-03-08) | Before |
| Cluster 2 | 2020-03-30 (2020-03-11; 2020-05-03) | 2020-03-18 (2020-03-11; 2020-03-26) | Undecided |
| Cluster 3 | 2020-03-25 (2020-03-15; 2020-04-12) | 2020-03-14 (2020-03-05; 2020-03-19) | Undecided |
| Cluster 4 | 2020-05-22 (2020-03-25; 2020-06-17) | 2020-03-31 (2020-03-22; 2020-04-07) | After |
| Cluster 5 | 2020-04-21 (2020-04-12; 2020-04-24) | 2020-03-30 (2020-03-19; 2020-04-07) | After |
| Cluster 6 | 2020-04-23 (2020-03-29; 2020-05-15) | 2020-03-22 (2020-03-07; 2020-04-20) | Undecided |
| Cluster 7 | 2020-07-04 (2020-06-08; 2020-07-10) | 2020-04-25 (2020-04-13; 2020-04-25) | After |

Table A1: TMRCAs and parent TMRCAs of clusters on a global tree.

*Exponential Growth Rate*

The estimates of the exponential growth rate $f$ have been produced using BEAST v1.10.4 together with the corresponding 95% uncertainty (i.e. highest posterior density interval or HPDI) intervals. The results are presented in Table A2.

| **Cluster** | **Model** | $\hat{f}$ **& 95% HPDI (Annual)** | $\hat{f}$ **& 95% HPDI (Daily)** |
|---|---|---|---|
| First | Exp. Growth | 19.206 (8.301; 30.011) | 0.052 (0.023; 0.082) |
| Second | Exp. Growth | 27.879 (6.499; 49.853) | 0.076 (0.018; 0.136) |
| Third | Exp. Growth | 28.050 (10.470; 48.294) | 0.077 (0.029; 0.132) |

Table A2: Estimates of exponential growth rate for the Ukrainian SARS-CoV-2 for the three largest clusters.
